# Effect of a sanitation intervention on the nutritional status of children in Maputo, Mozambique: a controlled before-and-after trial

**DOI:** 10.64898/2026.04.09.26350506

**Authors:** J Knee, T Sumner, Z Adriano, C Opondo, D Holcomb, E Viegas, R Nala, J Brown, O Cumming

## Abstract

**Background:** The rapid growth of the world’s urban population has contributed to the expansion of informal urban settlements in many cities across the world. In these settings, lack of safe sanitation combined with high population density and poverty contributes to heightened health risks for often vulnerable populations. The aim of this study was to evaluate the effect of a shared, onsite sanitation intervention on the nutritional status of children in Maputo, Mozambique.

**Methods:** The Maputo Sanitation (MapSan) trial was a controlled before-and-after study to evaluate the effect of a shared, onsite sanitation intervention on child health in Maputo, Mozambique. Here, we report the effects on childhood stunting, wasting and underweight, and height-for-age, weight-for-height and weight-for-age z-scores. Children were enrolled aged 1–48 months at baseline and outcomes were measured before and 12 and 24 months after the intervention, with concurrent measurement among children in a comparable control arm. The primary analysis was intention-to-treat. The trial was registered at ClinicalTrials.gov, number NCT02362932.

**Results:** We enrolled 757 and 852 children in the intervention and control groups respectively. There was no evidence for an effect of the intervention on any outcome at 12 or 24 months of follow-up except for wasting where there was very weak evidence for an effect (adjusted prevalence ratio: 0.497; 95% CI: 0.22-1.11; p=0.09). In two exploratory analyses - one including only those children born into compounds post-intervention and a second excluding children in control compounds which had independently improved their sanitation facilities during follow-up – we found that stunting increased in the intervention group whilst wasting decreased.

**Conclusions:** This study contributes to the growing evidence on the role of sanitation in shaping child health outcomes in informal urban settlements. We found no evidence for an effect on stunting and weak evidence for an effect on wasting. More research is needed to understand how sanitation can reduce childhood undernutrition in complex urban environments.

## INTRODUCTION

The rapid growth of the world’s urban population has contributed to the expansion of informal urban settlements in many cities across the world. Globally, the proportion of the urban population living in informal settlements slums has fallen slightly in recent decades from an estimated 28 % in 2000 to 25% in 2022. but the absolute number of people has risen (1). Over a billion people currently reside in informal settlements and this is expected to increase by a further 300 million people by 2030 (1). This growth presents numerous public health challenges as housing and infrastructure development fail to keep pace with the needs of the local population. Lack of adequate housing and public health infrastructure combined with high population density and poverty contributes to heightened health risks for often vulnerable populations (2).

Limited sanitation infrastructure in these complex urban environments is of particular concern. Sanitation – defined as the safe management of human waste – can prevent the transmission of a range of important enteric pathogens which cause diarrhoea and contribute to a large burden of morbidity and mortality globally, particularly among children (3). High population density as found in informal urban settlements likely compounds the risk associated with unsafe sanitation and increases the risk of disease transmission. Exposure to these enteric pathogens causes disease but is also associated with childhood undernutrition even when asymptomatic (4).

Observational studies in complex urban environments, including in Maputo, Mozambique, have reported a high burden of childhood undernutrition which is associated with poor sanitation (5). To date however, there have been no robust controlled intervention studies to directly assess the effect of improved sanitation on childhood undernutrition in informal settlements or slums. The Maputo Sanitation (MapSan) Trial was a controlled before-and-after (CBA) trial to evaluate the effect of a shared, onsite sanitation intervention in a dense urban environment on a range of child health outcomes (6). The research was conducted in informal, low-income neighbourhoods of Maputo, Mozambique, where high population density and inadequate sanitation contribute to a substantial burden of disease. We have previously reported the primary outcome of the trial - enteric pathogen detection in children using stool-based molecular methods (7) – as well as the effects on diarrhoea (5) mental health and sanitation-related quality of life (8). A cross-sectional analysis of the baseline data for this trial found that better quality sanitation was associated with reduced odds of stunting among children (5). The aim of this study was to assess the effect of the sanitation intervention on childhood stunting, wasting and underweight at twelve and twenty months of follow-up.

## METHODS

### Study Design & intervention

The MapSan trial was a controlled before-and-after (CBA) study that evaluated the impact of a shared onsite sanitation intervention on child health outcomes including enteric pathogen infection (stool-based enteric pathogen detection), diarrhoeal disease, and growth at 12- and 24-months post-intervention. Complete details of the trial design, intervention, and results of the primary outcome - enteric pathogen infection - have been published previously (6, 7).

The intervention was built by the non-governmental organisation (NGO), Water and Sanitation for the Urban Poor (WSUP) and consisted of shared pour-flush toilets connected to septic tank. Liquid effluent was discharged from the septic tank via a soakaway pit. Compounds with 20 or more residents received a communal sanitation block (CSB), which consisted of two or more toilet stalls (one per 20 residents), rainwater collection system, an elevated tank for storing municipal water, a stall for bathing, a laundry area, and a handwashing basin that could be connected to the municipal water supply or storage tank. Smaller compounds with fewer than 20 residents received a single stall shared toilet without the additional amenities included with the CSB. WSUP selected compounds for intervention based on a set of eight selection criteria: (1) existing shared sanitation facilities were of poor condition as determined by an engineer; (2) the compound was located in one of the 11 pre-defined implementation neighbourhoods; (3) there were a minimum of 12 residents; (4) residents were willing to contribute financially to construction costs; (5) there was sufficient space available for construction of the new facility; (6) the site was accessible for transportation of construction materials and tank-emptying activities; (7) the compound had access to a legal piped water supply; and (8) the groundwater level was deep enough for construction of a septic tank. Control compounds were selected by the study team based on a subset of the intervention site selection criteria including that residents shared sanitation in poor condition. Control compounds were also required to have at least one child <48 months old. Intervention site selection criteria related to facility construction and maintenance (criteria 5, 6, and 8) were not used for control selection. Due to a lack of eligible compounds in the 11 intervention neighbourhoods, control compounds were also selected from 5 adjacent and similar neighbourhoods. To the extent possible, control compounds were frequency matched with intervention compounds on resident number and time of enrolment.

### Enrolment & data collection

We enrolled children and collected data at baseline (pre-intervention) and at 12- and 24-months post-intervention. Children were eligible for enrolment if they were 1-48 months of age at baseline or would have been had they been present for baseline enrolment and if their parent or guardian provided written, informed consent. Children were ineligible if they had moved into the compound <6 months prior to the 12- or 24-month visit.

At each visit, we collected survey and observational data, including information on socio-demographics and environmental and WASH-related conditions and practices, in addition to outcome measures including child length/height and weight. Trained enumerators measured child height (or recumbent length, for children <2 years old) and weight in accordance with WHO standard methods (9). When possible, enumerators confirmed child birthdates using birth certificates.

### Outcomes

Our outcome measures were length/height-for-age z-score (HAZ), weight-for-age z-score (WAZ), and weight-for-height z-score (WHZ), which represent how child’s height or weight measurement compares with a healthy child of the same age and sex, as defined by the WHO Child Growth Standards (10). Children more than two standard deviations below the mean (<-2 z-score) were classified as stunted (HAZ), wasted (WHZ), and underweight (WAZ). We calculated and report HAZ, WAZ, and WHZ, and the binary variables stunted, underweight, and wasted, in accordance with the WHO and UNICEF guidelines and growth standards (10, 11) using the R package “anthro” (available at https://CRAN.R-project.org/package=anthro). We censored z-scores with implausible values, as recommended by the WHO guidelines for reporting growth data (11).

### Statistical analysis

We used a difference-in-difference analysis to assess the effect of the intervention on growth outcomes after 12- and 24-months. Difference-in-difference analyses estimate main effects as the interaction of the study arm and time-point of interest (baseline/12-month or baseline/24-month). We used generalised estimating equations (GEE) to fit linear regression models for continuous outcomes of HAZ, WAZ, and WHZ and estimate the mean difference in the change in z-score pre- and post-intervention between the intervention and control groups. We used GEE to fit Poisson regression models for binary outcomes to estimate prevalence ratios for stunting, underweight, and wasting in intervention versus control children after accounting for baseline differences and time effects. All GEE models included robust standard errors and accounted for clustering at the compound level. Multivariable models were adjusted for covariates determined *a priori* to be potential confounders, including child age, sex, breastfeeding status, household wealth score (12), and the primary caregiver’s education level. We assessed covariate balance across arms by calculating the standardised difference. A standardised difference of >0.1 indicated potential imbalance.

Our primary analysis included all children and used complete observations. We performed two additional exploratory analyses to assess the robustness of our results and variation among sub-groups. First, we estimated the intervention effects on children who were born into intervention and control compounds post-baseline, compared with children of a similar age at baseline, to assess the importance of exposure to the intervention from birth. Second, we estimated effects for children with longitudinal data, including only children who were present at both the baseline and 12-month time-points or at the baseline and 24-month time-points. The trial was originally designed to collect longitudinal measurements, however, due to population migration in and out of the study sites, our primary analysis treats data as repeated cross-sections. This longitudinal analysis also serves a sensitivity analysis to understand the potential impact of population migration on effect estimates in our primary analyses. Finally, as a sensitivity analysis, we estimated effects after excluding control compounds that independently upgraded their sanitation facilities to be similar to the WSUP intervention during the study period.

### Ethics

The study protocol was approved by the Comité Nacional de Bioética para a Saúde (CNBS), Ministério da Saúde (333/CNBS/14), the Research Ethics Committee of the London School of Hygiene & Tropical Medicine (reference # 8345), and the Institutional Review Board of the Georgia Institute of Technology (protocol # H15160). The trial is registered at ClinicalTrials.gov (NCT02362932).

## RESULTS

We enrolled 757 intervention children from 531 households in 235 compounds and 852 control children from 603 households in 297 compounds across all three time-points. Height/length or weight and birthdate data were available for 695 (91.8%) intervention children and 785 (92.1%) control children and of those children 3.45% (24/695 intervention) and 4.58% (36/785 control) of observations were excluded from analysis due to implausible z-scores (Table 1).

**Table 1:** Study population characteristics by arm at baseline.

|  | **Control** |  | **Intervention** |  |  |
| --- | --- | --- | --- | --- | --- |
|  | N | n (%) or mean (SD) | N | n (%) or mean (SD) | Standardized Difference<sup>1</sup> |
| **Child-level variables** |  |  |  |  |  |
| Age, days | 520 | 700 (405) | 441 | 694 (403) | 0.016 |
| Sex, female | 520 | 266 (51%) | 444 | 227 (51%) | 0.00055 |
| Breastfeeding, any | 526 | 169 (32%) | 448 | 143 (31%) | 0.0045 |
| Breastfeeding, exclusive | 526 | 49 (9%) | 448 | 37 (8%) | 0.037 |
| Child faeces disposed of in a latrine | 526 | 148 (28%) | 448 | 141 (31%) | 0.073 |
| Child wears diapers | 526 | 342 (65%) | 447 | 294 (65%) | 0.016 |
| Caregiver completed primary school | 528 | 287 (54%) | 451 | 239 (52%) | 0.027 |
| Child's mother is alive | 513 | 503 (98%) | 435 | 426 (97%) | 0.0085 |
| Respondent is child's mother | 519 | 368 (70%) | 443 | 284 (64%) | 0.15 |
| Stunted | 447 | 209 (46%) | 369 | 152 (41%) | 0.11 |
| Underweight | 459 | 90 (19%) | 379 | 71 (18%) | 0.022 |
| Wasted | 447 | 52 (11%) | 366 | 54 (14%) | 0.092 |
| Length/Height-for-age z-score | 447 | -1.7 (2.0) | 369 | -1.6 (1.9) | 0.058 |
| Weight-for-age z-score | 459 | -0.82 (1.4) | 379 | -0.81 (1.4) | 0.007 |
| Weight-for-height z-score | 447 | 0.19 (1.8) | 366 | 0.069 (1.8) | 0.07 |
| **Household-level variables** |  |  |  |  |  |
| Household population | 441 | 5.4 (2.4) | 365 | 6.1 (3.0) | 0.28 |
| Household wealth score, 0 (poorer) - 1 (wealthier)<sup>2</sup> | 440 | 0.45 (0.1) | 365 | 0.44 (0.1) | 0.15 |
| Crowding (Household member/room) | 440 | 2.3 (1.0) | 365 | 2.4 (1.2) | 0.085 |
| Household floor is covered<sup>3</sup> | 440 | 426 (96%) | 365 | 333 (91%) | 0.24 |
| Latrine used by household has a ceramic or masonry pedestal or slab<sup>3</sup> | 432 | 153 (35%) | 359 | 142 (39%) | 0.086 |
| Household drinking water source inside compound | 435 | 324 (74%) | 360 | 294 (81%) | 0.17 |
| **Compound-level variables** |  |  |  |  |  |
| Compound population | 287 | 14.1 (6.21) | 208 | 18.9 (12.1) | 0.50 |
| Number of households per compound | 287 | 3.8 (2.1) | 208 | 4.4 (3.7) | 0.19 |
| Number of water points per compound | 283 | 0.98 (0.95) | 207 | 1.4 (1.6) | 0.32 |
| Number of latrines per compound | 287 | 1.0 (0.2) | 207 | 1.1 (0.57) | 0.07 |
| Number of households sharing a latrine | 285 | 3.7 (1.8) | 197 | 4.0 (2.8) | 0.13 |
| Any animals present in compound<sup>3</sup> | 287 | 170 (59%) | 208 | 132 (63%) | 0.087 |

Data are n (%) or mean (standard deviation) and collected by questionnaire unless otherwise noted.
<sup>1</sup> Standardised difference of >0.1 suggests potential difference between arms.
<sup>2</sup> Assessed using Simple Poverty Scorecard for Mozambique (http://www.simplepovertyscorecard.com/MOZ_2008_ENG.pdf).
<sup>3</sup> Data collected by direct observation.

Most participant characteristics were balanced across arms across baseline, including age, sex, and breastfeeding status (Table 1). The child’s mother was more frequently the survey respondent for control children than intervention children at baseline. At the household level, intervention households had more residents on average and more frequently had access to a water source within the compound grounds, while control households tended to be slightly wealthier and more frequently had covered floors (a contributor to the wealth score). Intervention compounds were larger than control compounds, on average, with a higher average number of residents, households, and waterpoints. The average number of latrines per compounds was similar between intervention and controls, meaning that the average number of households sharing a latrine was higher among intervention compounds compared with controls.

At baseline, across both groups, the mean HAZ was -1.7 and 43.8% of children were classified as stunted (Figure 1, Table 2). While mean HAZ was similar among intervention and control children, the proportion of children stunted in intervention compounds (41%) was lower than in control children (47%). The mean WAZ and WHZ were similar across intervention and control children at baseline, as was the proportion underweight and wasted children.

**Figure 1:**
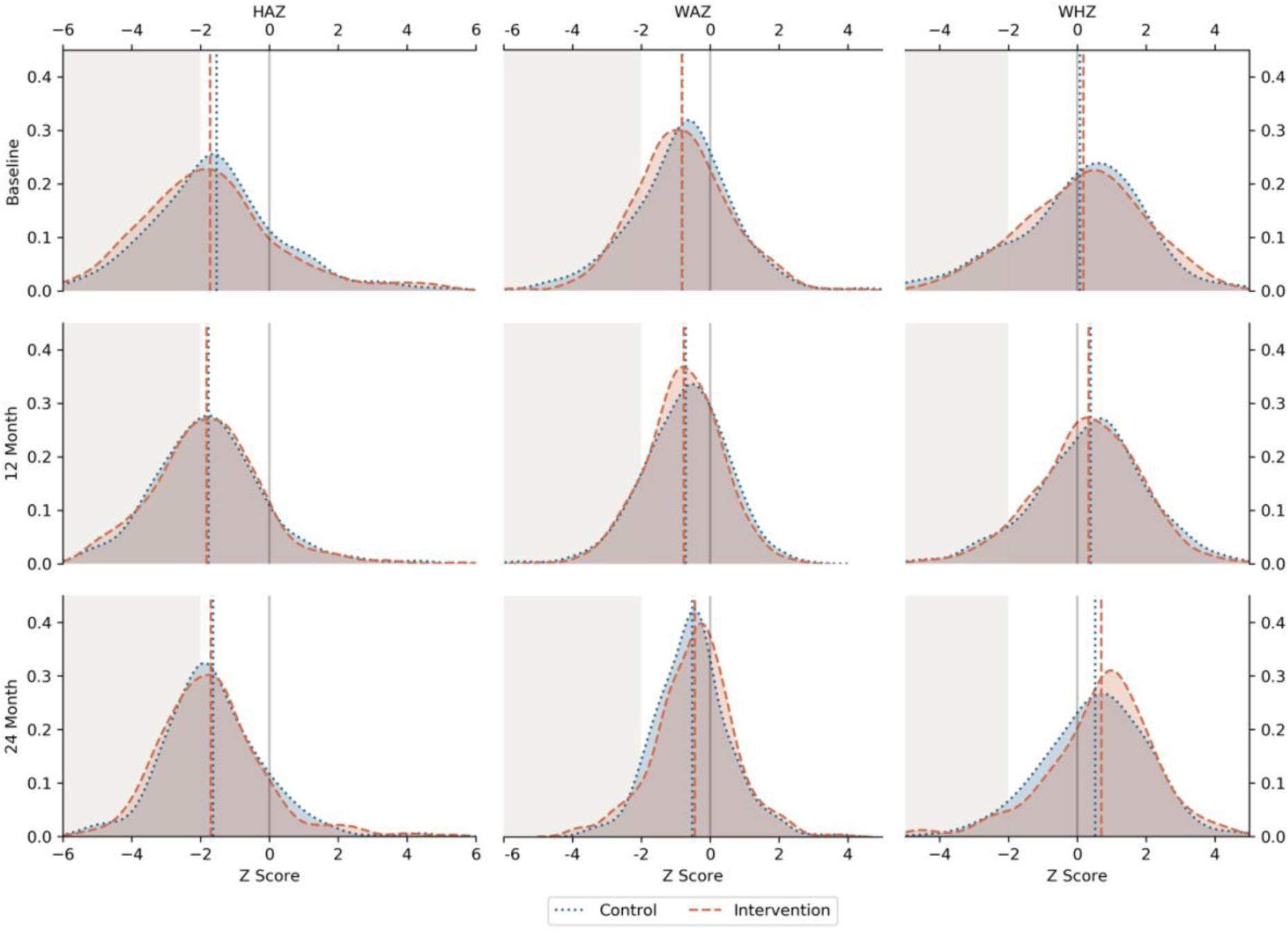
Kernel density plots for height-for-age (HAZ), weight-for-age (WAZ), and weight-for-height (WHZ) z-scores by arm at each study time-point (baseline, 12-month follow-up, 24-month follow-up). Vertical dashed lines represent the mean z-score for each arm. The grey shaded area represents the proportion of children stunted, underweight, and wasted.

**Table 2a:** Intervention effect on HAZ, WAZ, WFH at 12- and 24-month time-points among all childr.

|  | Baseline | 12-month | 24-month | 12-month effect estimate |  | 24-month effect estimate |  |
| --- | --- | --- | --- | --- | --- | --- | --- |
|  | Mean (SD) | Mean (SD) | Mean (SD) | Unadjusted | Adjusted | Unadjusted | Adjusted |
| **HAZ** |  |  |  |  |  |  |  |
| **control** | -1.69 (2.02)<br>n=447 | -1.84 (1.56)<br>n=410 | -1.66 (1.51)<br>n=309 |  |  |  |  |
| **Intervention** | -1.58 (1.85)<br>n=369 | -1.74 (1.56)<br>n=405 | -1.67 (1.42)<br>n=336 | -0.026 (-0.4, 0.35),<br>p=0.89 | -0.061 (-0.43, 0.3),<br>p=0.74 | -0.097 (-0.47, 0.27),<br>p=0.61 | -0.179 (-0.55, 0.19),<br>p=0.34 |
| **WAZ** |  |  |  |  |  |  |  |
| **control** | -0.82 (1.37)<br>n=459 | -0.77 (1.1)<br>n=412 | -0.45 (1.15)<br>n=310 |  |  |  |  |
| **Intervention** | -0.81 (1.44)<br>n=379 | -0.72 (1.22)<br>n=412 | -0.52 (1.07)<br>n=339 | 0.027 (-0.22, 0.28),<br>p=0.83 | -0.018 (-0.26,<br>0.22), p=0.88 | -0.066 (-0.33, 0.2),<br>p=0.63 | -0.087 (-0.34, 0.16),<br>p=0.5 |
| **WHZ** |  |  |  |  |  |  |  |
| **control** | 0.19 (1.76)<br>n=447 | 0.35 (1.49)<br>n=409 | 0.68 (1.55)<br>n=308 |  |  |  |  |
| **Intervention** | 0.07 (1.81)<br>n=366 | 0.37 (1.61)<br>n=406 | 0.55 (1.5)<br>n=337 | 0.15 (-0.19, 0.49),<br>p=0.39 | 0.136 (-0.2, 0.48),<br>p=0.43 | 0.001 (-0.37, 0.37),<br>p=0.99 | 0.077 (-0.29, 0.44),<br>p=0.68 |

**Table 2b:** Intervention effect on stunting, underweight, and wasting at 12- and 24-month time-points among all children.

|  | Baseline | 12-month | 24-month | 12-month prevalence ratio, 95% CI |  | 24-month prevalence ratio, 95% CI |  |
| --- | --- | --- | --- | --- | --- | --- | --- |
|  | % (95 CI), n | % (95 CI), n | % (95 CI), n | Unadjusted | Adjusted | Unadjusted | Adjusted |
| **Stunted** |  |  |  |  |  |  |  |
| <u>control</u> | 0.47 (0.42 - 0.51)<br>n=447 | 0.45 (0.4 - 0.5)<br>n=410 | 0.43 (0.37 - 0.48)<br>n=309 |  |  |  |  |
| <u>Intervention</u> | 0.41 (0.36 - 0.46)<br>n=369 | 0.43 (0.39 - 0.48)<br>n=405 | 0.41 (0.36 - 0.47)<br>n=336 | 1.094 (0.87,<br>1.38), p=0.45 | 1.106 (0.88,<br>1.39), p=0.39 | 1.089 (0.85,<br>1.39), p=0.49 | 1.135 (0.89,<br>1.45), p=0.31 |
| **Underweight** |  |  |  |  |  |  |  |
| <u>control</u> | 0.19 (0.16 - 0.23)<br>n=459 | 0.13 (0.1 - 0.17)<br>n=412 | 0.1 (0.07 - 0.14)<br>n=310 |  |  |  |  |
| <u>Intervention</u> | 0.18 (0.15 - 0.23)<br>n=379 | 0.15 (0.11 - 0.18)<br>n=412 | 0.06 (0.04 - 0.1)<br>n=339 | 1.162 (0.75,<br>1.8), p=0.5 | 1.228 (0.79,<br>1.9), p=0.36 | 0.723 (0.39,<br>1.36), p=0.31 | 0.76 (0.41,<br>1.42), p=0.39 |
| **Wasted** |  |  |  |  |  |  |  |
| <u>control</u> | 0.11 (0.09 - 0.15)<br>n=447 | 0.07 (0.05 - 0.1)<br>n=409 | 0.06 (0.04 - 0.1)<br>n=308 |  |  |  |  |
| <u>Intervention</u> | 0.14 (0.11 - 0.18)<br>n=366 | 0.08 (0.05 - 0.11)<br>n=406 | 0.05 (0.03 - 0.08)<br>n=337 | 0.892 (0.48,<br>1.67), p=0.72 | 0.887 (0.47,<br>1.67), p=0.71 | 0.578 (0.26,<br>1.27), p=0.17 | 0.497 (0.22,<br>1.11), p=0.09 |

Our primary analysis included all children enrolled in the study who had valid z-scores. We found no evidence for an effect of the intervention on HAZ, WHZ, WAZ, stunting or underweight at 12- and 24-months in adjusted or unadjusted analyses (Tables 2a,2b). We found very weak evidence for an effect on wasting (aPR: 0.497; 95% CI: 0.22-1.11; p=0.09). While the proportion of underweight and wasted children appeared to decrease over time (at each time-point) in both groups, there appeared to be a larger decrease in the proportion wasted in the intervention group, with prevalence ratios below the null value of 1.0 for all analyses, though none were statistically significant (Figure 2, Table 2b). The proportion of stunted children in each group was comparatively static across time-points (Figure 2).

**Figure 2:**
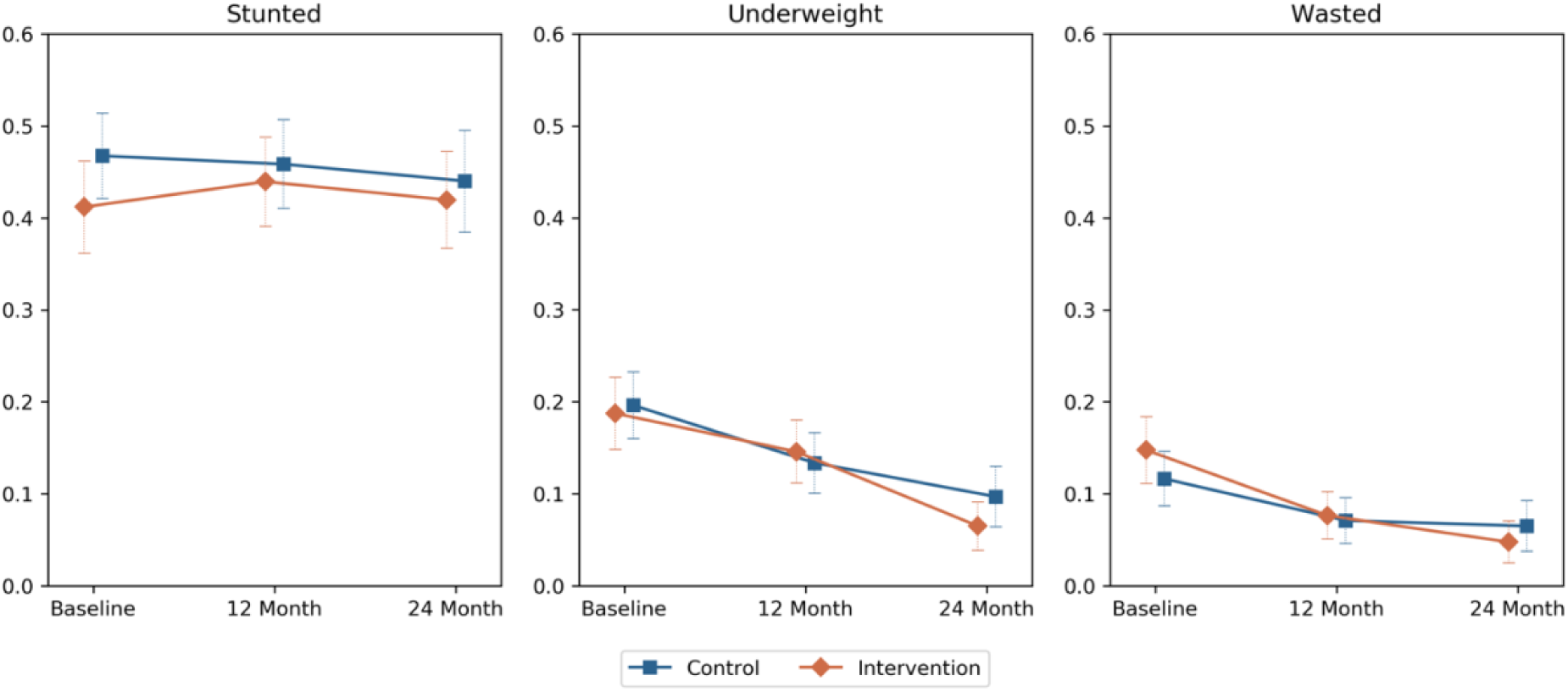
Proportion of children stunted, underweight, and wasted in intervention and control arms at baseline, 12-month, and 24-months post-intervention.

We assessed the effect of having the intervention from birth via a sub-group analysis which included only children who were born into the intervention (or control) compounds post-baseline but before the 12- or 24-month time-point, compared with children of a similar age group at baseline. Few children met these criteria at 12-months of follow-up, and we found no evidence for an effect of the intervention on any outcome at 12 months of follow-up (Table 3a), though the precision of our effect estimates was low with very wide confidence intervals. At 24 months of follow-up, the mean difference in the change in HAZ between intervention and control children in adjusted analyses was -0.63 (95% confidence interval (CI): -1.26 – 0.00, p=0.051) suggesting HAZ worsened among intervention children compared with control. We found a similar effect at 24-months for stunting, with a larger increase in stunting prevalence in intervention children compared with control (adjusted prevalence ratio (aPR) 1.596, 95% CI 1.08 – 2.35, p=0.02, Table 3b). Conversely, we observed a reduction in the prevalence of wasting among intervention children compared with controls after controlling for differences at baseline and changes over time (aPR 0.182, 95% CI 0.04 – 0.81, p=0.03). This was consistent with the results for WHZ (Table 3a), which had a mean difference of 0.52 (95% CI: -0.04 – 1.08, p=0.07) although evidence for this was weak. The intervention had no effect on WAZ or the binary outcome underweight.

**Table 3a:** Intervention effect on HAZ, WAZ, WFH among children born into intervention compounds by the 12- or 24-month time-point compared with children of a similar age at baseline.

|  | Baseline ≤1 years | 12-month, Born-in & ≤1 year | 12-month effect estimate |  | Baseline ≤2 years | 24-month, Born-in & ≤2 years | 24-month effect estimate |  |
| --- | --- | --- | --- | --- | --- | --- | --- | --- |
|  | Mean (SD) | Mean (SD) | Unadjusted | Adjusted | Mean (SD) | Mean (SD) | Unadjusted | Adjusted |
| **HAZ** |  |  |  |  |  |  |  |  |
| control | -0.87 (2.4)<br>n=118 | -1.34 (2.04)<br>n=50 |  |  | -1.39 (2.31)<br>n=247 | -1.61 (1.77)<br>n=101 |  |  |
| Intervention | -0.81 (2.27)<br>n=91 | -1.25 (2.26)<br>n=65 | 0.033 (-1.02, 1.08), p=0.95 | 0.079 (-0.95, 1.1), p=0.88 | -1.16 (2.1)<br>n=193 | -1.74 (1.7)<br>n=98 | -0.384 (-1.04, 0.27), p=0.25 | -0.629 (-1.26, 0.00), p=0.051 |
| **WAZ** |  |  |  |  |  |  |  |  |
| control | -0.17 (1.54)<br>n=124 | -0.32 (1.15)<br>n=50 |  |  | -0.49 (1.42)<br>n=254 | -0.22 (1.19)<br>n=101 |  |  |
| Intervention | -0.28 (1.62)<br>n=97 | -0.11 (1.23)<br>n=69 | 0.325 (-0.27, 0.92), p=0.29 | 0.381 (-0.23, 0.99), p=0.22 | -0.4 (1.44)<br>n=201 | -0.1 (1.1) n=98 | 0.026 (-0.36, 0.42), p=0.9 | -0.047 (-0.45, 0.35), p=0.82 |
| **WHZ** |  |  |  |  |  |  |  |  |
| control | 0.66 (2.06)<br>n=114 | 0.85 (2.05)<br>n=50 |  |  | 0.42 (1.85)<br>n=243 | 0.81 (1.72)<br>n=98 |  |  |
| Intervention | 0.19 (2.06)<br>n=89 | 1.01 (1.67)<br>n=63 | 0.632 (-0.23, 1.49), p=0.15 | 0.652 (-0.19, 1.49), p=0.13 | 0.27 (1.91)<br>n=190 | 1.06 (1.63)<br>n=97 | 0.433 (-0.14, 1), p=0.14 | 0.52 (-0.04, 1.08), p=0.07 |

**Table 3b:** Intervention effect on stunting, underweight, and wasting among children born into intervention compounds by the 12- or 24-month time-point compared with children of a similar age at baseline.

|  | **Baseline ≤1**<br>**years** | **12-month,**<br>**Born-in & ≤1**<br>**year** | **12-month** |  | **Baseline ≤2**<br>**years** | **24-month,**<br>**Born-in & ≤2**<br>**years** | **24-month** |  |
| --- | --- | --- | --- | --- | --- | --- | --- | --- |
|  | % (95% CI),<br>n | % (95% CI), n | Unadjusted | Adjusted | % (95% CI), n | % (95% CI), n | Unadjusted<br>Prevalence<br>Ratio, 95% | Adjusted<br>Prevalence<br>Ratio |
| **Stunted** |  |  |  |  |  |  |  |  |
| <u>control</u> | 0.36 (0.27 -<br>0.45) n=118 | 0.42 (0.28 -<br>0.57) n=50 |  |  | 0.45 (0.38 -<br>0.51) n=247 | 0.45 (0.35 -<br>0.55) n=101 |  |  |
| <u>Intervention</u> | 0.31 (0.22 -<br>0.41) n=91 | 0.45 (0.32 -<br>0.57) n=65 | 1.221 (0.67,<br>2.23), p=0.52 | 1.229 (0.68,<br>2.23), p=0.50 | 0.33 (0.27 -<br>0.4) n=193 | 0.49 (0.39 -<br>0.59) n=98 | 1.472 (1,<br>2.16), p=0.05 | 1.596 (1.08,<br>2.35), p=0.02 |
| **Underweight** |  |  |  |  |  |  |  |  |
| <u>control</u> | 0.11 (0.06 -<br>0.18) n=124 | 0.08 (0.02 -<br>0.19) n=50 |  |  | 0.16 (0.11 -<br>0.21) n=254 | 0.06 (0.02 -<br>0.12) n=101 |  |  |
| <u>Intervention</u> | 0.11 (0.06 -<br>0.19) n=97 | 0.06 (0.02 -<br>0.14) n=69 | 0.724 (0.16,<br>3.24), p=0.67 | 0.667 (0.15,<br>2.94), p=0.59 | 0.11 (0.07 -<br>0.17) n=201 | 0.03 (0.01 -<br>0.09) n=98 | 0.743 (0.19,<br>2.86), p=0.67 | 0.773 (0.2,<br>2.96), p=0.71 |
| **Wasted** |  |  |  |  |  |  |  |  |
| <u>control</u> | 0.13 (0.08 -<br>0.21) n=114 | 0.12 (0.05 -<br>0.24) n=50 |  |  | 0.12 (0.08 -<br>0.17) n=243 | 0.08 (0.04 -<br>0.15) n=98 |  |  |
| <u>Intervention</u> | 0.16 (0.09 -<br>0.25) n=89 | 0.05 (0.01 -<br>0.13) n=63 | 1.202 (0.61,<br>2.37), p=0.60 | 0.349 (0.08,<br>1.47), p=0.15 | 0.14 (0.09 -<br>0.19) n=190 | 0.03 (0.01 -<br>0.09) n=97 | 0.31 (0.08,<br>1.15), p=0.08 | 0.182 (0.04,<br>0.81), p=0.03 |

We assessed the impact of the intervention among children with repeated measures at baseline and 12-months and baseline and 24-months. Results were consistent with the primary analysis and the intervention had no effect on any of the growth outcomes in this sub-group analysis (Table 4a, 4b).

**Table 4a:** Intervention effect on HAZ, WAZ, WFH among children with complete data at Baseline & 12-month time-points and at Baseline & 24-month time-points.

|  | Baseline | 12-month | 12-month |  | Baseline | 24-month | 24-month |  |
| --- | --- | --- | --- | --- | --- | --- | --- | --- |
|  | Mean (SD) | Mean (SD) | Unadjusted | Adjusted | Mean (SD) | Mean (SD) | Unadjusted | Adjusted |
| **HAZ** |  |  |  |  |  |  |  |  |
| <u>control</u> | -1.74 (1.93)<br>n=230 | -1.83 (1.43)<br>n=231* |  |  | -1.68 (2.06)<br>n=125 | -1.64 (1.5)<br>n=125 |  |  |
| <u>Intervention</u> | -1.61 (1.94)<br>n=220 | -1.68 (1.36)<br>n=219 | 0.004 (-0.43,<br>0.44), p=0.98 | -0.015 (-0.44,<br>0.41), p=0.95 | -1.51 (2)<br>n=141 | -1.53 (1.16)<br>n=141 | -0.067 (-0.62,<br>0.49), p=0.81 | -0.157 (-0.69,<br>0.38), p=0.57 |
| **WAZ** |  |  |  |  |  |  |  |  |
| <u>control</u> | -0.76 (1.23)<br>n=237 | -0.74 (1.05)<br>n=237 |  |  | -0.73 (1.22)<br>n=127 | -0.69 (1.09)<br>n=127 |  |  |
| <u>Intervention</u> | -0.76 (1.31)<br>n=224 | -0.78 (1.19)<br>n=224 | -0.04 (-0.31,<br>0.23), p=0.77 | -0.027 (-0.3,<br>0.24), p=0.84 | -0.67 (1.26)<br>n=144 | -0.75 (0.95)<br>n=144 | -0.126 (-0.46,<br>0.2), p=0.46 | -0.185 (-0.51,<br>0.14), p=0.27 |
| **WFH** |  |  |  |  |  |  |  |  |
| <u>control</u> | 0.3 (1.74)<br>n=225 | 0.32 (1.33)<br>n=225 |  |  | 0.24 (1.73)<br>n=122 | 0.4 (1.44)<br>n=122 |  |  |
| <u>Intervention</u> | 0.08 (1.77)<br>n=215 | 0.24 (1.57)<br>n=215 | 0.138 (-0.29,<br>0.57), p=0.53 | 0.154 (-0.28,<br>0.59), p=0.49 | 0.21 (1.87)<br>n=140 | 0.23 (1.34)<br>n=140 | -0.037 (-0.52,<br>0.45), p=0.88 | -0.138 (-0.7,<br>0.42), p=0.63 |

**Table 4b:** Intervention effect on stunting, underweight, and wasting among children with complete data at Baseline & 12-month time-points and at Baseline & 24-month time-points.

|  | Baseline | 12-month | 12-month |  | Baseline | 24-month | 24-month |  |
| --- | --- | --- | --- | --- | --- | --- | --- | --- |
|  | % (95% CI), n | % (95% CI), n | Unadjusted | Adjusted |  | % (95% CI), n | Unadjusted | Adjusted |
| **Stunted** |  |  |  |  |  |  |  |  |
| <u>control</u> | 0.47 (0.4 - 0.54) n=230 | 0.42 (0.36 - 0.49) n=231* |  |  | 0.47 (0.38 - 0.56) n=125 | 0.42 (0.33 - 0.51) n=125 |  |  |
| <u>Intervention</u> | 0.42 (0.36 - 0.49) n=220 | 0.38 (0.32 - 0.45) n=219 | 1.004 (0.75, 1.35), p=0.98 | 1.034 (0.78, 1.38), p=0.82 | 0.4 (0.32 - 0.48) n=141 | 0.35 (0.27 - 0.43) n=141 | 1.043 (0.58, 1.86), p=0.89 | 1.057 (0.74, 1.51), p=0.76 |
| **Underweight** |  |  |  |  |  |  |  |  |
| <u>control</u> | 0.15 (0.11 - 0.2) n=237 | 0.13 (0.09 - 0.18) n=237 |  |  | 0.14 (0.09 - 0.21) n=127 | 0.13 (0.07 - 0.2) n=127 |  |  |
| <u>Intervention</u> | 0.16 (0.12 - 0.22) n=224 | 0.15 (0.1 - 0.2) n=224 | 1.072 (0.59, 1.93), p=0.82 | 1.043 (0.58, 1.86), p=0.89 | 0.13 (0.08 - 0.2) n=144 | 0.08 (0.04 - 0.13) n=144 | 0.933 (0.37, 2.36), p=0.88 | 0.707 (0.28, 1.78), p=0.46 |
| **Wasted** |  |  |  |  |  |  |  |  |
| <u>control</u> | 0.09 (0.06 - 0.13) n=225 | 0.05 (0.02 - 0.09) n=225 |  |  | 0.1 (0.05 - 0.17) n=122 | 0.07 (0.03 - 0.13) n=122 |  |  |
| <u>Intervention</u> | 0.14 (0.1 - 0.2) n=215 | 0.07 (0.04 - 0.12) n=215 | 0.944 (0.37, 2.38), p=0.9 | 0.933 (0.37, 2.36), p=0.88 | 0.13 (0.08 - 0.2) n=140 | 0.06 (0.02 - 0.11) n=140 | 0.992 (0.69, 1.43), p=0.97 | 0.588 (0.17, 2.07), p=0.41 |

Between baseline and the 12-month visits, 17 control compounds independently upgraded their sanitation facilities, with a further 15 upgrading by the 24-month visit. We conducted a sensitivity analysis excluding children living in control compounds with upgraded sanitation facilities, and the results were largely consistent with the primary analysis. The intervention had no effect on any growth outcome with the exception of wasting at 24-months, where the intervention was associated with a 55% reduction in prevalence compared with control children after controlling for baseline differences and changes over time (aPR 0.445, 95% CI 0.20 – 0.99, p=0.05) (Table 5).

**Table 5:** Intervention effect on HAZ, WAZ, WFH and binary outcomes stunting, underweight, and wasting among children living in intervention compounds or control compounds that did <u>not</u> upgrade their sanitation during the study period compared with results from the primary analysis.

|  |  | 12-month |  |  | 24-month |  |
| --- | --- | --- | --- | --- | --- | --- |
|  | n | Unadjusted | Adjusted* | n | Unadjusted | Adjusted |
| **HAZ** |  |  |  |  |  |  |
| <u>Primary analysis (including all compounds)</u> | 1631 | -0.026 (-0.4, 0.35),<br>p=0.89 | -0.061 (-0.43, 0.3),<br>p=0.74 | 1461 | -0.097 (-0.47, 0.27),<br>p=0.61 | -0.179 (-0.55, 0.19),<br>p=0.34 |
| <u>Excluding upgraded sanitation</u> | 1610 | -0.026 (-0.41, 0.35),<br>p=0.89 | -0.063 (-0.43, 0.31),<br>p=0.74 | 1432 | -0.107 (-0.49, 0.27),<br>p=0.58 | -0.217 (-0.59, 0.16),<br>p=0.26 |
| **WAZ** |  |  |  |  |  |  |
| <u>Primary analysis (including all compounds)</u> | 1662 | 0.027 (-0.22, 0.28),<br>p=0.83 | -0.018 (-0.26, 0.22),<br>p=0.88 | 1487 | -0.066 (-0.33, 0.2),<br>p=0.63 | -0.087 (-0.34, 0.16),<br>p=0.5 |
| <u>Excluding upgraded sanitation</u> | 1641 | 0.025 (-0.23, 0.28),<br>p=0.85, n=1641 | -0.016 (-0.26, 0.23),<br>p=0.9 | 1458 | -0.019 (-0.29, 0.25),<br>p=0.89 | -0.048 (-0.31, 0.21),<br>p=0.72 |
| **WHZ** |  |  |  |  |  |  |
| <u>Primary analysis (including all compounds)</u> | 1628 | 0.15 (-0.19, 0.49),<br>p=0.39 | 0.136 (-0.2, 0.48),<br>p=0.43 | 1458 | 0.001 (-0.37, 0.37),<br>p=0.99 | 0.077 (-0.29, 0.44),<br>p=0.68 |
| <u>Excluding upgraded sanitation</u> | 1607 | 0.148 (-0.19, 0.49),<br>p=0.4, n=1607 | 0.141 (-0.2, 0.48),<br>p=0.42 | 1430 | 0.057 (-0.33, 0.44),<br>p=0.77 | 0.146 (-0.23, 0.52),<br>p=0.45 |
| **Stunted** |  |  |  |  |  |  |
| <u>Primary analysis (including all compounds)</u> | 1631 | 1.094 (0.87, 1.38),<br>p=0.45 | 1.106 (0.88, 1.39),<br>p=0.39 | 1461 | 1.089 (0.85, 1.39),<br>p=0.49 | 1.135 (0.89, 1.45),<br>p=0.31 |
| <u>Excluding upgraded sanitation</u> | 1610 | 1.099 (0.87, 1.39),<br>p=0.43 | 1.114 (0.88, 1.4),<br>p=0.36 | 1432 | 1.091 (0.85, 1.4),<br>p=0.49 | 1.152 (0.9, 1.48),<br>p=0.27 |
| **Underweight** |  |  |  |  |  |  |
| <u>Primary analysis (including all compounds)</u> | 1662 | 1.162 (0.75, 1.8),<br>p=0.50 | 1.228 (0.79, 1.9),<br>p=0.36 | 1487 | 0.723 (0.39, 1.36),<br>p=0.31 | 0.76 (0.41, 1.42),<br>p=0.39 |
| <u>Excluding upgraded sanitation</u> | 1641 | 1.177 (0.76, 1.84),<br>p=0.47 | 1.236 (0.79, 1.93),<br>p=0.35 | 1458 | 0.656 (0.35, 1.23),<br>p=0.19 | 0.691 (0.37, 1.29),<br>p=0.25 |
| **Wasted** |  |  |  |  |  |  |
| <u>Primary analysis (including all compounds)</u> | 1628 | 0.892 (0.48, 1.67),<br>p=0.72 | 0.887 (0.47, 1.67),<br>p=0.71 | 1458 | 0.578 (0.26, 1.27),<br>p=0.17 | 0.497 (0.22, 1.11),<br>p=0.09 |
| <u>Excluding upgraded sanitation</u> | 1607 | 0.879 (0.47, 1.65),<br>p=0.69 | 0.875 (0.46, 1.66),<br>p=0.68 | 1430 | 0.525 (0.24, 1.16),<br>p=0.11 | 0.445 (0.2, 0.99),<br>p=0.05 |

\*n for adjusted analyses slightly smaller than listed in table due to missing covariate data.

## DISCUSSION

We conducted a controlled before-and-after trial in informal urban neighbourhoods of Maputo, Mozambique to estimate the effect of a sanitation intervention on undernutrition among children aged 6-59 months at 12 and 24 months of follow-up. There are two main findings. Firstly, in the primary analysis, there was no evidence for an effect of the intervention on any outcome at 12 or 24 months of follow-up except for wasting where there was very weak evidence for an effect. Secondly, in two exploratory analyses - one including only those children born into compounds post-intervention and a second excluding children in control compounds which had independently improved their sanitation facilities during follow-up – we found that stunting increased in the intervention group whilst wasting decreased.

There was no evidence that the sanitation intervention improved height-for-age, weight-for-height or height-for-age z-scores nor reduced the prevalence of stunting or underweight at either 12 or 24 months of follow-up. There was weak evidence for an effect of the intervention on the prevalence of wasting at 24 months of follow-up (aPR: 0.497; 95% CI: 0.22-1.11; p=0.09). The proportion of children stunted was high in this population but similar in the two arms at 12 and 24 months of follow-up time (43% and 41% respectively in intervention compounds and 45% and 43% in control compounds). These findings are consistent with the previously reported results for the main outcomes of the MapSan trial; there was no evidence for an effect of this intervention on enteric infection, soil-transmitted re-infection nor diarrhoea (7). Our findings are broadly consistent with findings from various recent trials of sanitation interventions albeit in rural areas which reported no effect of sanitation on linear growth (13–15). There are a number of distinguishing factors between these previous trials and our study, though. Firstly, the type of sanitation evaluated. In the previous trials conducted in rural areas the sanitation intervention consisted of basic pit latrines whereas in our study the sanitation intervention comprised pour-flush toilets discharging to septic tanks with the liquid effluent released through a soakaway pit (7). In addition, the sanitation facilities in our study were shared by multiple households, unlike in the previous trials where facilities were not shared (7). Secondly, the urban setting of our study – relatively high density informal urban settlements - was quite different to the rural settings of the previous studies. Stunting is a chronic multifactorial condition reflecting sustained nutritional deficits and cumulative disease history requiring sustained comprehensive strategies for its prevention. Furthermore, in this densely populated urban setting children were inevitably exposed directly or indirectly to hazards beyond the compounds — through shared outdoor spaces, open drains, and contact with individuals from outside of the compound.

We conducted two secondary exploratory analyses: one where we restricted our analysis to children who were born after the intervention was delivered and another where we excluded children from control compounds that independently improved their sanitation facilities during follow-up. Under both of these analyses, we found evidence that at 24 months of follow-up the prevalence of stunting increased in the intervention arm compared to the control but conversely the prevalence of wasting decreased in the intervention arm compared to the control. The findings in these two secondary analyses with regard to a reduction in wasting at 24 months are consistent with the weak evidence for an effect in the primary analysis. When the analysis was restricted to only children born after the intervention had been delivered there was weak evidence of an effect on WHZ (difference-in-difference of 0.52 WHZ; 95% CI: -0.04-1.08; p-value 0.07), and evidence of an effect on wasting (aPR: 0.497; 95% CI: 0.22-1.11; p=0.09). Notably when the control compounds which had independently improved their sanitation facility during follow-up – which would bias estimates towards the null - were excluded there was stronger evidence for an effect (aPR: 0.445; 95% CI 0.20–0.99, p=0.05). Unlike stunting, which reflects cumulative long-term nutritional deficits, wasting is an indicator of acute undernutrition and may be more immediately sensitive improvements in environmental conditions. Reductions in enteric pathogen exposure — as previously reported in this trial — could plausibly improve nutrient absorption and reduce the acute metabolic demands of infection, thereby supporting weight gain in young children. The consistent directionality of the wasting estimates across the primary, longitudinal, and sensitivity analyses lends tentative biological plausibility to this signal, even if it falls short of statistical certainty. The contamination of the control arm through independent sanitation upgrades in 32 compounds by 24 months is likely to have attenuated the estimated intervention effect, and the strengthening of the wasting effect estimate after excluding these compounds supports this interpretation. Previous trials have reported no effect on wasting but as discussed above these concerned settings and the sanitation interventions that were very different to our study (13–15). Larger trials with greater statistical power would be needed to detect changes of the magnitude observed in our study without statistical evidence.

This study has several limitations that should be considered when interpreting the results. First, the intervention was not randomly allocated and neither participants nor the research team could be blinded to the allocation status, which may introduce potential selection bias, and observer bias with regard to outcome assessment (16). Second, there was some contamination that occurred between study arms as certain control compounds independently improved their sanitation during the period of follow-up; however, sensitivity analyses were conducted to assess the impact of these upgrades on the findings. Third, the intervention focused on compound-level sanitation improvements rather than community-wide sanitation coverage, which may limit the overall potential reduction in environmental faecal contamination and associated health risks such as stunting. In addition, the generalisability of the findings may be limited because living conditions and sanitation infrastructure can vary widely across complex urban environments, meaning results from these neighbourhoods in Maputo, Mozambique may not be representative of other complex urban settings.

In conclusion, this study contributes to the growing evidence on the role of sanitation in shaping child health outcomes in informal urban settlements. We found no evidence for an effect on stunting and weak evidence for an effect on wasting. More research is needed to understand how sanitation can reduce childhood undernutrition in complex urban environments.

## Data Availability

All data produced in the present study are available upon reasonable request to the author

## ACKNOWLEDGEMENTS

We gratefully acknowledge data collection services and other support provided by the WE Consult team and in particular Wouter Rhebergen and Ellen de Bruijn, and the hard work of the enumerators Isabel Maninha Chiquele, Sergio Adriano Macumbe, Carolina Zavale, Maria Celina Macuacua, Guil-herme Zimba, and Anabela Mondlane, as well as Olimpio Zavale who coordinated logistics. We thank staff at the Instituto Nacional de Saude, specifically Josina Mate, Acacio Sabonete, and Jeronimo Langa, for their support throughout the project.

The study would not have been possible without Water and Sanitation for the Urban Poor (WSUP), in particular Guy Norman, Carla Costa, Vasco Parente, and Sam Drabble. We also thank our advisory group for their technical input in the design of the study: Ben Arnold, Jack Colford, Jay Graham, Tony Kolb, Eddy Perez, Tom Slaymaker, Larry Moulton, and Darren Saywell. We also thank Radu Ban from the Gates Foundation for his support and advice throughout the project. Finally, and most importantly of all, we thank all of the participants and their families who graciously welcomed us into their homes and were so generous with their time.

